# Evaluating a Large Reasoning Model’s Performance on Open-Ended Medical Scenarios

**DOI:** 10.1101/2025.04.29.25326666

**Authors:** R. E. Hoyt, D. Knight, M. Haider, M. Bajwa

**Affiliations:** Virginia Commonwealth University, Richmond, VA; Mayo Clinic Jacksonville, FL; Carilion Clinic, Roanoke, VA; MGH Institute of Health Professions, Boston, MA

## Abstract

Large language models (LLMs) have emerged as a dominant form of generative artificial intelligence (GenAI) in multiple domains. In early 2025, DeepSeek R1 was released, which is a new large reasoning model (LRM) that includes CoT (CoT) reasoning, Mixture of Experts (MoE), and reinforcement learning. As these technologies continue to improve, evaluating the accuracy and reliability of LLMs and LRMs in medicine remains a crucial challenge. This paper reports on a follow-up study using DeepSeek R1 to evaluate medical scenarios contained in the MMLU-Pro benchmark, an enhanced benchmark designed to evaluate language understanding models across broader and more challenging tasks. In the previously reported study, the accuracy rate was 96% when multiple-choice MMLU-Pro answers were provided. In the current study, we evaluated DeepSeek R1 on 162 medical scenarios, but without multiple-choice answers provided. The overall accuracy was 92%. This approach mirrors a more realistic clinical scenario where the clinician must decide on the most likely diagnosis and differential diagnoses without any clues. Further research is necessary to determine how to deploy LRMs in clinical medicine, given their high accuracy rate, both with and without answers provided.

## Introduction

This study builds on our previous evaluation of the DeepSeek R1 large reasoning model (LRM), analyzing complex medical scenarios from the MMLU-Pro benchmark. [1-3] Our earlier work analyzed DeepSeek R1’s accuracy in correctly identifying the correct answers for 162 medical scenario-related questions, covering 21 specialties. This medical closed-ended question bank had four to ten possible answers for each medical scenario. Subject matter experts (SMEs) reviewed and confirmed the correct answers for twenty-three questions where DeepSeek’s response differed from the provided answer, determining the overall accuracy to be 96%. [1] According to SMEs, sixty-five percent of DeepSeek R1’s answers were correct when they disagreed with MMLU-Pro’s answers. The mean number of references cited by DeepSeek was 16 (range 0-46), and the mean number of reasoning steps was 12.2 (range 3-56). No hallucinated references were noted, although unrelated references were common. In the earlier study we did not see any correlation between incorrect answers and the number of references or reasoning steps. We concluded that DeepSeek R1 was highly accurate with closed-ended questions that involved medical knowledge and reasoning skills. We also noted that this LRM was among the most transparent by including multiple references and reasoning steps.

While our initial research provided valuable insights into DeepSeek R1’s ability to select the correct answer from a given set of options, limitations inherent in multiple-choice questions (MCQs) warrant further investigation. As noted in the original manuscript, multiple-choice questions can suffer from the “cueing effect” and “testwiseness,” potentially allowing test-takers, or a large language model, to arrive at the correct answer without deep reasoning. [4-5] Furthermore, MCQs may not adequately assess the ability to understand and apply knowledge in complex, real-world situations, often focusing more on factual recall than complex clinical reasoning.

In this subsequent phase of our research, we aim to provide a more valid evaluation of DeepSeek R1’s capabilities in analyzing complex medical scenarios. To achieve this, we employed the same set of 162 medical scenarios previously utilized, but this time, we presented the questions to DeepSeek R1 without providing any multiple-choice answer options (close-ended questions). [6] By taking away cues for test-taking strategies, this study aims to give a more realistic evaluation of DeepSeek R1’s ability to analyze medical information on its own. We evaluated its capacity to generate potential diagnoses and articulate its reasoning, free from the constraints of given answer choices. The results of this investigation will further contribute to our understanding of how LRMs like DeepSeek R1 can be leveraged in the complex domain of clinical medicine.

## Methods

The MMLU-Pro question bank, which consists of about 12,000 questions and answers in 14 categories, was downloaded from the hosting website using Python programming. [7] The health category was selected by filtering, and this category consisted of the following subcategories: virology, clinical knowledge, anatomy, medical genetics, nutrition, college medicine, aging, and professional medicine. We further filtered this category to only include the professional medicine subcategory, which included 166 complex medical scenarios. Questions were further categorized into twenty-one medical specialties. Approximately a third of the questions were related to adult or pediatric primary care. About 15% were emergency room visits, and the remainder were divided among multiple subspecialties.

In our earlier study, we became aware that DeepSeek R1 disagreed with the MMLU-Pro answer on 23 occasions. We decided to submit the same questions to Claude 3.7 Sonnet Reasoning as a second opinion, and in a majority of cases it agreed with DeepSeek R1. We therefore submitted these questions to subject matter experts (SMEs), who agreed with DeepSeek R1 on 15 questions and disagreed on 8 questions. We established these answers as “ground truth” before testing MMLU-Pro medical scenarios with no answers supplied.

We deleted duplicate questions and deleted four question and answer pairs that contained errors. We created a prompt that stated, “*As a senior clinician, you have received requests for consultation on various medical scenarios. Provide a diagnosis and differential diagnosis*.”

We used a well-known AI-enabled search engine that hosted an uncensored version of DeepSeek R1, as well as two other LRMs, o3 mini and Claude 3.7 Sonnet Reasoning. [8-9] We inputted the prompt and the scenario in a single paragraph. For a majority of scenarios, the last sentence was changed from “Which of the following is most likely…” to “What is the most likely….” DeepSeek R1 was selected from a list of LLMs and LRMs, as well as the option to select academic papers. DeepSeek R1 performed the following actions: 1. It first performed a literature review to gather relevant information. 2. It began an iterative process that consisted of multiple steps as part of CoT reasoning. 3. It then provided a diagnosis and differential diagnosis. Lastly, the answer was followed by an explanation of why it was chosen and why it excluded the other potential answers, often supported by literature citations. (See an example of the DeepSeek R1 workflow in Appendix A.) In the Appendix, the three steps above are displayed as a typical workflow. The reasoning steps and key diagnostic considerations incorporate resource citations.

## Results

One hundred and sixty-two question/answer pairs were analyzed with DeepSeek R1. There was agreement in 149 cases, or 92% accuracy, which is less than the 96.3% accuracy noted in our previous study of the same question/answer pairs when multiple-choice answers were included.

References, unrelated references, and reasoning steps were calculated for the 162 scenarios, and the results are displayed in Table 1. In 19 scenarios there were no references included, and in 95 scenarios there were no unrelated references. There was one unrelated reference outlier with 70 unrelated references. The question addressed a statistical issue in an HIV patient, with the majority of references pertaining to HIV rather than statistics.

**Table 1.** References, unrelated references and reasoning steps.

| Category | Minimum | Maximum | Average |
| --- | --- | --- | --- |
| References | 0 | 84 | 33 |
| Unrelated references | 0 | 70 | 3 |
| Reasoning steps | 3 | 34 | 11 |

As stated in our earlier paper, we are unable to explain why DeepSeek R1 included no references for certain scenarios, and we were also unable to explain the variability in unrelated references. [1] In 59% (95/162), there were no unrelated references. Furthermore, as we found in our earlier work, the related and the unrelated references were real and not hallucinated.

### Analyzing the Errors

In the current study, thirteen errors were identified. It should be stressed that this study required more thought and scrutiny than the original study, as DeepSeek R1 occasionally generated correct or appropriate answers not listed among the MMLU-Pro answers. For example, a college student had a close exposure to a classmate with meningococcal meningitis, but the only relevant answer for treatment listed by MMLU-Pro was rifampin. In this study, DeepSeek R1 suggested ceftriaxone, which is an appropriate alternative and therefore a correct answer, even though it was not mentioned as an MMLU-Pro choice. For any scenario initially deemed incorrect, three clinicians reviewed all references and reasoning steps. In one instance, a more recent answer contradicted the MMLU-Pro answer, which was based on a 2008 guideline. In our original article and in this study, we identified multiple MMLU-Pro answers that were incorrect, outdated, or had reasonable alternatives. There have been multiple studies that tested LLMs and LRMs on MedQA-type questions, but it is unclear how often clinician authors confirmed the validity of the answers. The 13 “incorrect” answers were reviewed and adjudicated by three board-certified internal medicine physicians (RH, MH, and DK). Table 2 displays the scenarios where the DeepSeek answer did not align with the MLU-Pro answer, followed by a discussion and the results of retesting. Table definitions are as follows:

**Table 2.** DeepSeek vs MMLU-Pro answers, error discussion, and retest results.

| Scenario Number:<br>DeepSeek Answer | MMLU-Pro Answer | Error Discussion | Test-Retest Results |
| --- | --- | --- | --- |
| 6118: Hand arm vibration syndrome (HAVS) | Axillary-subclavian venous thrombosis | Anchoring bias: The patient presented with acute symptoms and arm swelling as evidence against HAVS. Factual error: the answer is not listed as an MMLU-Pro answer | Incorrect |
| 6181: Hyperviscosity syndrome | Atlanto-axial instability | Anchoring bias: Focused on facial plethora. Confirmation bias: the answer is not listed as an MMLU-Pro answer | Correct |
| 6236: Anthrax | Corynebacterium diphtheriae | Factual error: DeepSeek mentioned corynebacterium in its reasoning but stated it's less common, which is not true. The final differential diagnosis did not include corynebacterium. | Incorrect |
| 6242: STD. Treat with triple antibiotics. Herpes simplex proctitis | Imiquimod for anal warts. | Anchoring bias: DeepSeek did mention HPV condylomata in differential diagnoses. It is true that anal warts rarely cause pain. | Correct |
| 6359: Peptic ulcer disease | Herpes zoster | Factual error and anchoring bias: It was stated that the lack of a rash reduced the likelihood of a diagnosis, although pain frequently precedes the rash. | Incorrect |
| 6379: Thrombolytic agent | Nitroglycerin increasing nitric oxide concentration | Factual error: stated "Since the pain is recurring, perhaps they're considering nitro for symptom relief, but nitro doesn't address the clot." The question asked about immediate treatment. | Correct |
| 6435: Herpes simplex | Chancroid | Anchoring bias: the vaginal fluid smear could be helpful for both HSV and chancroid. | Correct |
| 6542: Urine albumin to creatinine ratio (uACR) | Flu shot | Factual error: DeepSeek failed to refer to the 2007 NKF or 2025 ADA guidelines. According to the guidelines, checking uACR should begin 5 years after the diagnosis of type I diabetes. The ADA guidelines were included in the references. | Incorrect |
| 6664: Egg donation | Adopt a child | Indeterminate answer: The SME recommended genetic counseling. This case is an unusual scenario where a 13-year-old girl with Turner's syndrome wants to have babies. | Incorrect |
| 6672: decreased lymphocyte proliferative response to mitogens | Am cortisol | Factual error and confirmation bias: DeepSeek did mention the elevated cortisol level during stress but failed to mention testing it. | Incorrect |
| 6674: Pre-menstrual dysphoria syndrome | Major depressive disorder | Factual error, anchoring and confirmation biases: symptoms were more chronic than PMDD. | Incorrect |
| 6748: Respiratory tract or skin portal, hematogenous spread. | Olfactory nerve | Factual error and anchoring bias: question focused on amebas other than N. fowleri. One of its references confirmed that it enters the brain via the olfactory nerve. | Incorrect |
| 6793: Recommended upper endoscopy | Urine pregnancy test | Anchoring bias: DeepSeek concentrated on blood-tinged vomit rather than more common etiologies for her age group | Incorrect |

- Incorrect: The answer deviates from what we determined as the ground truth based on the SME output.
- Indeterminate: The answer was inconclusive based on included facts.
- Anchoring bias: LRM chose one concept and followed that one CoT.
- Confirmation bias: The tendency to favor information that confirms existing beliefs while ignoring or dismissing contradictory information.
- Factual error: The information is incorrect and does not align with established and verifiable evidence.

For question 6435, the original diagnosis did not agree with the MMLU-Pro answer. We repeated the same scenario for DeepSeek R1 because other LRMs concurred with the MMLU-Pro response. On repeat testing, DeepSeek R1 changed its answer to align with MMLU-Pro. We proceeded to retest all incorrect answers, and in four cases, DeepSeek R1 changed the answer to align with MMLU-Pro (see Table 2). This finding demonstrates that DeepSeek R1 has reduced test-retest reliability, although it appears to be infrequent. In a study reported by Mykhalko et al., they did perform retesting using four LLMs and noted that the retest accuracy was 97% for ChatGPT 4o and Claude 3.5 Sonnet. [10] We are unaware of any studies that evaluated test-retest reliability in LRMs.

It is of interest that none of the errors noted in our earlier study were the same errors noted in this study. We also explored whether the references cited and the reasoning steps were any different between the correct and incorrect answers. An unpaired Student t-test (performed in Google Sheets) revealed no statistical difference between the average number of references (p = 0.510), but there was a statistical difference in the average number of reasoning steps (p = 0.028). (See Table 3). The increased reasoning steps suggest that when DeepSeek R1 was in doubt, it required more reasoning steps and evidence of “overthinking” for some scenarios. The small number of incorrect reasoning steps limits statistical testing, however.

**Table 3.** References and Reasoning Steps.

| Category | Correct | Incorrect | p-values |
| --- | --- | --- | --- |
| References | 33 | 36 | 0.510 |
| Reasoning steps | 11 | 15 | 0.028 |

## Discussion

The diagnostic accuracy on MMLU-Pro medical scenarios was excellent and almost as good as our original study, which demonstrated an accuracy of 96.3%. Improved diagnostic accuracy is important for inpatient and outpatient medical care, particularly for reducing preventable errors. According to the National Academy of Sciences, inpatient diagnostic errors are responsible for 6 to 17 percent of adverse events and more closed malpractice claims than other causes. [11]

Our study was not the first to test LLMs on open-ended medical scenarios. In 2023, Kung et al. reported the results of analyzing 376 USMLE questions with ChatGPT 3.5 using a closed, and open-ended approach. The accuracy for the open-ended approach was the lowest (45%) on USMLE Step 1, which focused on basic science, and better (65%) on Step 3, which focused on clinical knowledge. Surprisingly, about 40% of answers were deemed “indeterminate.” [12] Nachane et al. analyzed 500 MEDQA-USMLE open-ended question/answer pairs with Llama-7B and Llama-70B. The accuracy achieved was 83% and 87%, respectively. [13] MyKhalko et al. reviewed 100 medical cases of unknown origin. Phase I involved open-ended questions with four LLMs: Gemini, GPT 3.5, GPT 4o, and Claude 3.5 Sonnet. Accuracy for open-ended questions varied from 44 to 72%. In contrast, the accuracy ranged from 65 to 95% for closed-ended questions. [10] We could not find a study that submitted open-ended questions to DeepSeek R1 or another LRM.

There have been multiple studies reported in 2025 that evaluated DeepSeek R1 on close-ended medical MCQs, and all demonstrated high accuracy. Moell et al. used qualitative and quantitative analyses of 100 diverse clinical cases from the MedQA dataset and demonstrated that DeepSeek R1 achieved a 93% accuracy rate, similar to our results. They evaluated each error for trends and reported various mistakes and biases made by the LRM. [14]

Mikhail et al. evaluated 300 ophthalmology cases from StatPearls using DeepSeek R1 and OpenAI o1. Both had an accuracy rate of 82%. They emphasized that there was a 15-fold reduction in query per token expenses with DeepSeek R1 compared to OpenAI o1. [15] In another published study on ophthalmology cases, 130 multiple-choice questions (MCQs) related to diagnosis and management were collected from the Chinese ophthalmology senior professional title examination and translated into English. DeepSeek R1, Gemini 2.0 Pro, OpenAI o1, and OpenAI o3-mini attained accuracies of 0.808, 0.746, 0.723, and 0.577 in English MCQs, respectively. [16]

Mondillo et al. evaluated 500 pediatric MCQs from the MEDQA dataset using DeepSeek R1 and OpenAI o1. DeepSeek scored lower at 87%, and OpenAI’s accuracy was 92.8%. [17] In another study by Mondillo et al., they analyzed 900 pediatric MCQs from the MEDQA dataset using the LRMs o3 mini and o3 mini high. They both scored well with accuracies of 88.3% and 90.5%, respectively. They introduced the notion that perhaps an ensemble of more than one LRM would be even more accurate. [18]

Another study evaluated the performance of DeepSeek-R1 in diagnosing oral diseases using text-based cases from the New England Journal of Medicine’s “Image Challenge” series that included answer choices. DeepSeek-R1 achieved a diagnostic accuracy of 91.6 %, greatly exceeding the 47.8 % average accuracy of journal readers. [19]

One of the perceived advantages of LRMs is that they display their reasoning steps. Such information represents new transparency; however, there is preliminary evidence that the thinking steps are not necessary for excellent LRM accuracy. [20] Furthermore, in a 2025 study by Anthropic, they concluded that the reasoning steps do not reflect what actually happens in a model. The model may determine the answer first and then simply add the reasoning steps later. [21] Therefore, we don’t know if LRMs are really any more accurate for analyzing medical scenarios than frontier LLMs and if they are really more transparent. We postulate that the overthinking phenomenon manifests in several recurring patterns: 1) Analysis paralysis: models become trapped in extended reasoning chains without reaching actionable conclusions. 2) Rogue actions: models take premature or inappropriate actions after overthinking. 3) Premature disengagement: models give up after extensive but unproductive reasoning. In other words, do these patterns lead the model to just use its first impression?

An area of research that needs further exploration is the addition of retrieval augmented generation (RAG), where the LRM has access to external resources. There is ample evidence that the RAG methodology results in improved LLM accuracy. [22]

## Limitations of LLMs and LRMs

Recent information suggests that LLMs and LRMs score well on standard medical MCQs and can score better than physicians. [23] LRMs score well on medical knowledge and reasoning, but expert and effective medical care requires even more skills. Knowing when to treat and when not to treat is important, as is including the patient’s and family’s treatment wishes. While LLMs and LRMs can express empathy, they cannot detect non-verbal cues related to common life experiences or interject humor when appropriate. LLMs and LRMs cannot provide a physical touch, and they can’t deliver bad news effectively, which requires empathy, timing, situational awareness, and the ability to manage strong emotional responses.

There are other challenges associated with LLMs and LRMs worth noting. Work by Omar et al. has shown that fabricated information included in a prompt may cause the model to hallucinate misleading information. The study tested DeepSeek R1 and other LLMs on 300 medical cases, altering one fact in each prompt. The hallucination rate for DeepSeek was very high and raises the concern that inadvertent or intentional “adversarial attacks” on the LLM or LRM could have very significant consequences. [24] LRMs have also recently been investigated for “faithfulness,” or how well the steps outlined in the CoT reasoning steps actually correspond to actions by the model. Preliminary studies suggest that DeepSeek R1 has a low level of unfaithfulness. [25-26] Furthermore, the recency of training data for DeepSeek R1 is unknown, and it failed safety checks. [27] For those reasons, healthcare organizations may be reluctant to use this LRM.

While many posit that LLMs and LRMs augment our intelligence, there is concern about the impact of these technologies on critical thinking in the field of medicine. Generative AI will save time by cognitive offloading, but there is qualitative and quantitative evidence that they reduce critical thinking. [28-29] Medical schools have the daunting task of finding a “sweet spot” for teaching the right amount of generative AI at the right time with the right patient. [30]

## Limitations and future implications

This study analyzed only one LRM and one medical MCQ-related dataset. Results might be different for other medical scenarios and LRMs. We also did not experiment with prompting. We chose a basic prompt, and one shortcoming we noted was that we did not ask the LRM for a single diagnosis. On several occasions, it produced the same answer as noted by MMLU-Pro, but a reasonable alternative as well. Studying the unpredictable results of changing the prompt is essential. There is no guarantee that creating a more complex prompt, particularly with LRMs, will result in higher accuracy. [31] Additionally, we couldn’t modify the DeepSeek R1’s temperature and could only surmise that it was in the low range.

It is unlikely that a single LLM or LRM will be 100% accurate with questions that cover so many specialties. The addition of RAG, combining two or more LLMs or LRMs, or model fine-tuning will likely improve the accuracy, but it will also increase model complexity. Further research is needed in this area.

Given their excellent accuracy but known limitations, the more crucial question is how to use evolving frontier LLMs and LRMs in clinical medicine. Our original article outlined the need for more research. It also stressed that these models should be tested with newer, more expansive, and challenging medical benchmarks before they are deployed in the field of medicine. [32-33]

## Conclusion

DeepSeek R1 has excellent accuracy in evaluating complex medical scenarios with and without answers provided. From an educational or research perspective, new LRMs are more transparent and easier to evaluate compared to new frontier LLMs. Further research is needed to determine if LRMs are really more accurate, particularly as newer LLMs continue to evolve with more features such as larger context windows and multimodal capabilities.

## Data Availability

All data is available on FigShare

https://www.figshare.com

## Notes

1. Quillbot was used to detect spelling and syntax errors
2. The MMLU-Pro MCQs and spreadsheets for the original study and the second study are available on FigShare (https://figshare.com) Log in and search for the MMLU-Pro Project

Appendix A

The prompt comes first, followed by the scenario number. Note the 30 sources and the 17 tasks (reasoning steps)

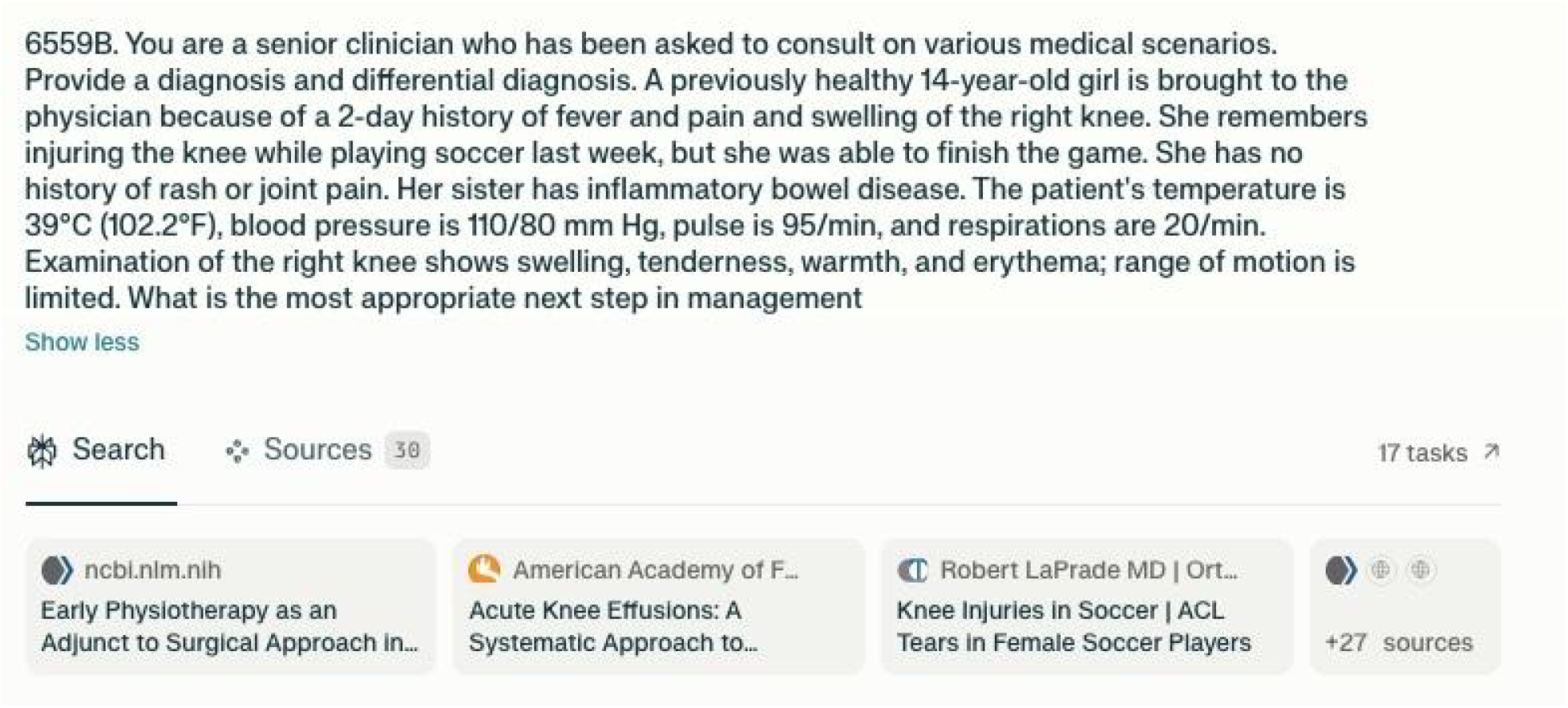

Reasoning Steps

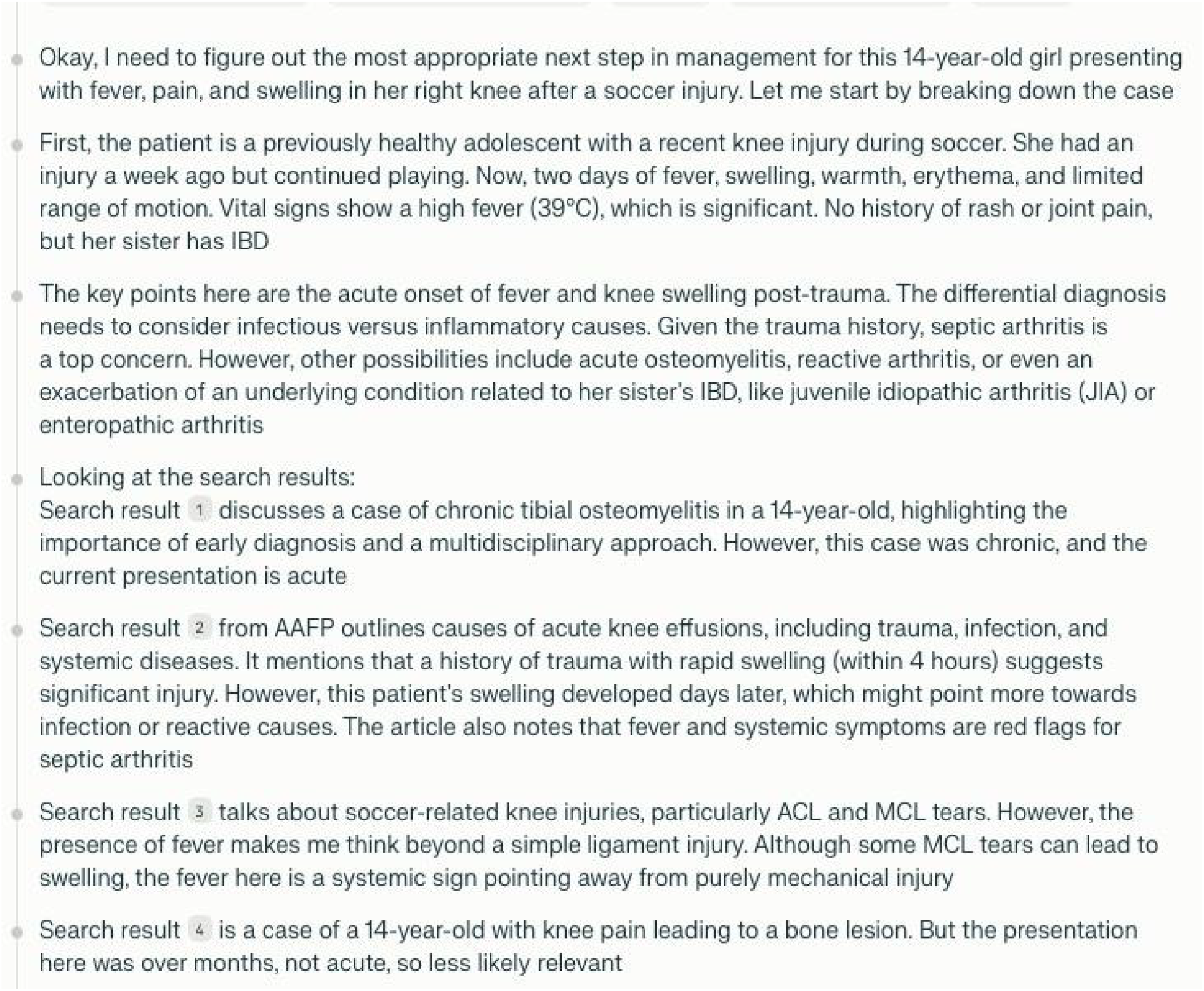

Diagnosis and Differential Diagnosis

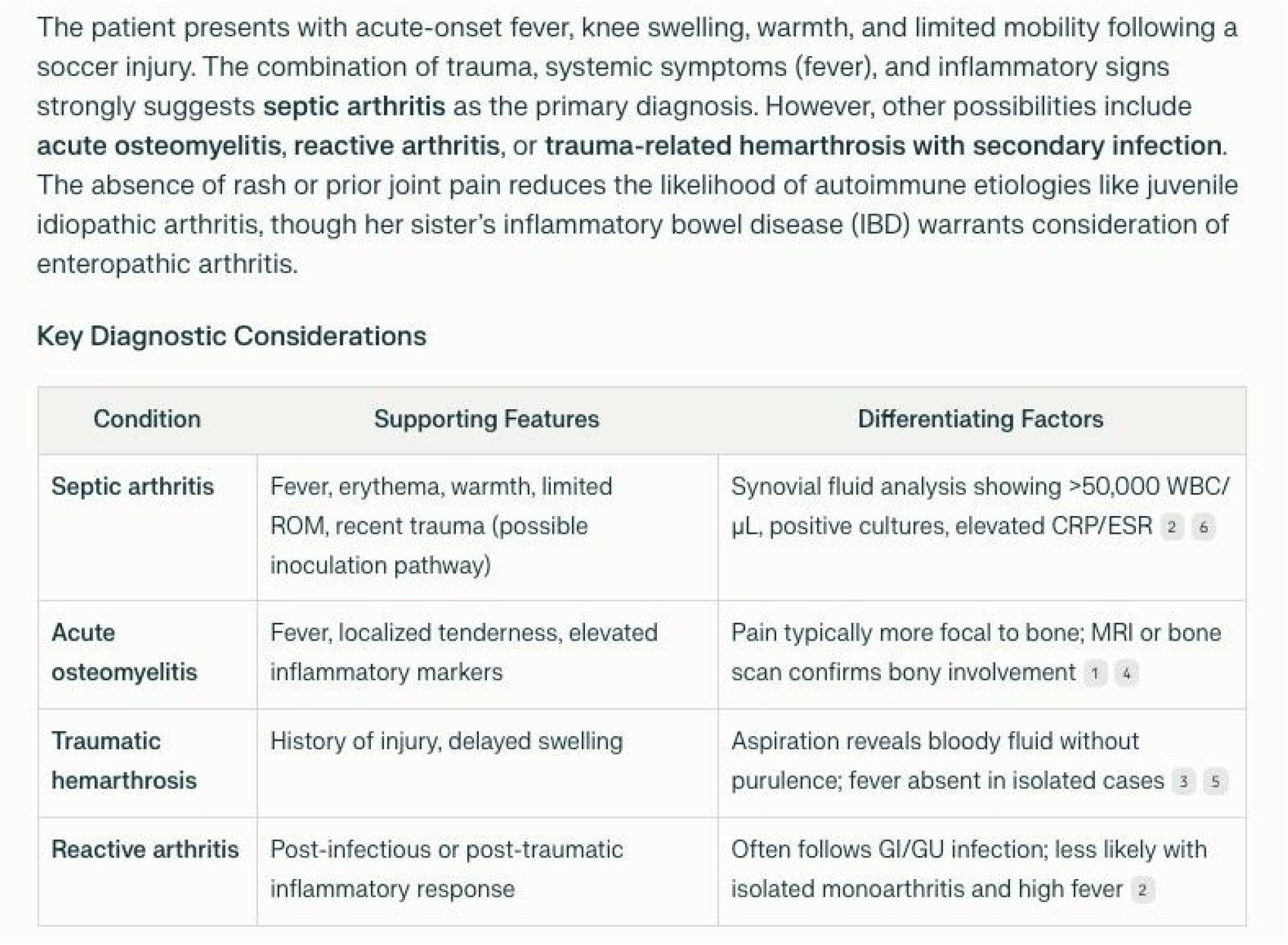

